# The PKN1- TRAF1 signaling axis as a potential new target for chronic lymphocytic leukemia

**DOI:** 10.1101/2021.03.22.21253655

**Authors:** Maria I. Edilova, Jaclyn C. Law, Safoura Zangiabadi, Kenneth Ting, Achire N. Mbanwi, Andrea Arruda, David Uehling, Methvin Isaac, Michael Prakesch, Rima Al-awar, Mark D. Minden, Ali A. Abdul-Sater, Tania H. Watts

## Abstract

TRAF1 is a pro-survival adaptor molecule in TNFR superfamily (TNFRSF) signaling. TRAF1 is overexpressed in many B cell cancers including refractory chronic lymphocytic leukemia (CLL). Little has been done to assess the role of TRAF1 in human cancer. Here we show that the protein kinase C related kinase Protein Kinase N1 (PKN1) is required to protect TRAF1 from cIAP-mediated degradation during constitutive CD40 signaling in lymphoma. We show that the active phospho-Thr774 form of PKN1 is constitutively expressed in CLL but minimally detected in unstimulated healthy donor B cells. Through a screen of 700 kinase inhibitors, we identified two inhibitors, OTSSP167 and XL-228, that inhibited PKN1 in the nanomolar range and induced dose-dependent loss of TRAF1 in RAJI cells. OTSSP167 and XL-228 treatment of primary patient CLL samples led to a reduction in TRAF1, pNF-κB p65, pS6, pERK, Mcl-1 and Bcl-2 proteins, and induction of activated caspase-3. OTSSP167 synergized with venetoclax in inducing CLL death, correlating with loss of TRAF1, Mcl-1 and Bcl-2. Although correlative, these findings suggest the PKN1-TRAF1 signaling axis as a potential new target for CLL. These findings also suggest OTSSP167 and venetoclax as a combination treatment for TRAF1 high CLL.

## Introduction

CLL is the most common human leukemia with an estimated 21,040 new cases and 4060 deaths in the US in 2020 (https://seer.cancer.gov/statfacts/html/clyl.html). CLL is considered to be largely a disease of the lymph node and bone marrow, where CLL cells receive survival signals through their B cell receptor (BCR).^1, 2^ Several promising new therapies for CLL target BCR signaling or cell survival, including the phosphatidyl inositol-3-kinase inhibitor idelalisib, the Bruton’s tyrosine kinase (BTK) inhibitor ibrutinib, as well as the Bcl-2 antagonist venetoclax.^3-8^ While these treatments are showing great promise, responses are not always durable and when relapse occurs there are limited treatment options available.^3, 9, 10^

In addition to signaling through the BCR, B cells require signals through TNFRSF members such as CD40, which activates NF-κB to induce pro-survival Bcl-2 family members.^11^ Many human malignancies including CLL, B cell lineage non-Hodgkins lymphoma and Burkitt’s lymphomas, exhibit constitutive signaling via TNFRs, such as CD40, CD30 or the EBV protein LMP1.^12-14^ TNFRSF members trigger NF-κB activation through recruitment of TRAF proteins.^15^ TRAF1 is an NF-κB inducible protein^16^ that on its own cannot induce NF-κB but as a 1:2 heterotrimer with TRAF2 recruits a single cellular inhibitor of apoptosis protein (cIAP)^17^ to induce activation of the classical NF-κB signaling pathway downstream of several TNFRSF members, including CD40 on B cells.^18^ cIAPs are dual function E3 ligases. cIAPs can add K63-linked polyubiquitin to RIP, thereby activating NF-κB signaling,^19^ but can also add K48-linked ubiquitin to TRAF proteins, thereby inducing TRAF protein degradation.^20^ Thus an outstanding question is how TRAF proteins can recruit cIAPs without being degraded. TRAF1 is overexpressed in 48% of B cell related cancers with the highest expression in the most refractory CLL.^21^ Non-coding single nucleotide polymorphisms in *TRAF1* have been linked to non-Hodgkin’s lymphoma;^22^ however, TRAF1 dependent signaling has not been specifically targeted as a cancer therapy. While TRAF2, the binding partner for TRAF1 is ubiquitously expressed and mice deficient in TRAF2 die of lethal inflammation,^23^ TRAF1 expression is largely limited to cells of the immune system^24^ and mice lacking TRAF1 have no overt phenotype.^25^ The finding that *Traf1* is required for lymphomagenesis in a spontaneous mouse tumor model induced by constitutively active NF-κB2^26^ makes TRAF1 a potential target in B cell malignancies; however, signaling adaptors are not readily amenable to drug discovery.

TRAF1 is a target of phosphorylation by the ubiquitously expressed protein kinase C related kinase PKN1 (also known as PRK1).^27^ PKN1 phosphorylates TRAF1, but not other TRAF family members, at serine 146 in human or serine 139 in mouse.^27^ Here we show that phosphorylation of TRAF1 S146 by PKN1 is required to protect TRAF1 from cIAP-mediated degradation during constitutive CD40 signaling in RAJI cells. We find that the activated phospho-Thr744 form of PKN1 is readily detected in patient CLL cells but minimally present in B cells from healthy donors. Accordingly, we screened a library of 700 kinase inhibitors and identified two PKN1 inhibitors, OTSSP167, a maternal embryonic leucine zipper kinase (MELK) inhibitor and XL-228, a multi-targeted tyrosine kinase inhibitor, that induced dose-dependent decreases in TRAF1, NF-κB p65, pERK, pS6, Mcl-1 and Bcl-2 proteins in primary patient CLL cells, with concomitant induction of cell death. We also report that OTSSP167 has synergistic effects with the Bcl-2 antagonist venetoclax in inducing cell death.

## Methods

### Human subjects

Frozen peripheral blood mononuclear cells (PBMCs) from 25 CLL patients were obtained from the Leukemia Tissue Bank at Princess Margaret Cancer Centre/University Health Network (UHN), Toronto, Ontario (**Table S1**). Informed consent for tissue bank donation was in compliance with the Declaration of Helsinki and in agreement with the UHN Research Ethics Review Board (protocol 01-0573) and the research herein was also approved by the University of Toronto Research Ethics board (Protocol #: 00030910). Cytogenetic alterations in CLL samples were determined by fluorescent *in situ* hybridization. Researchers were blinded to the clinical status of the patient samples until study completion. CLL cells were identified as CD19^+^/CD5^+^ cells by flow cytometry and generally accounted for >85% of PBMC. Blood from healthy donors was obtained with informed consent (University of Toronto REB protocol number 00027673) and samples stored as frozen PBMC until use. Healthy donor B cells were purified from PBMC using the EasySep Human B cell Isolation Kit (STEMCELL Technologies) according to the manufacturer’s protocol.

### Cell lines

RAJI and Daudi cell lines were from ATCC, 293 cells from in Invitrogen. For further details on cell culture and procedures see supplemental methods. The human embryonic kidney 293 cell line (293 FT) was obtained from Invitrogen (Invitrogen, Carlsbad, United States) and cultured in Dulbecco’s modified Eagle’s medium (Sigma-Aldrich, St. Louis, United States) supplemented with 10% Fetal Calf Serum (FCS, Thermo Fisher Scientific) and 1% of 100X Glutamine-Penicillin-Streptomycin (GPPS) (Sigma-Aldrich, Oakville, Canada). RAJI and Daudi were obtained from the ATTC and maintained in RPMI 1640 with FCS/GPPS. All cell lines tested negative for mycoplasma (Mycoplasma detection kit, Millipore-Sigma, Oakville, Ontario). OP9 cells were kindly provided by J.C. Zuniga-Pflücker, Sunnybrook research institute, Toronto, Canada and plated 10^5^ cells/well of a 24-well plate in α-MEM supplemented with 20% FCS and GPPS and used when confluent.

### Plasmid construction and transfection

The human TRAF1 ORF was amplified from RAJI cells cDNA, cloned into pENTR-D-TOPO vector (Invitrogen) and the S146A mutation introduced into pENTR-D-TOPO using QuikChange II Site-Directed Mutagenesis Kit (Stratagene, San Diego, United States) following manufacturer’s instructions. WT-TRAF1 and S146A-TRAF1 were cloned into a pcDNA3-c-Flag vector, a gift from Stephen Smale (Addgene plasmid # 20011). The Flag tag in WT-TRAF1 construct was replaced with a 3x-HA tag. WT and K644E human PKN1 ORFs were codon optimized for mammalian gene expression and synthesized by GeneArt Gene Synthesis (Invitrogen). WT and K644E PKN1 were the cloned into pCDNA3.1+ (Invitrogen). Cells were transfected with either 200ng of WT-TRAF1-HA, 200ng of S146A-TRAF1-FLAG, 50ng of WT PKN1, 50ng of K644E-PKN1 or 50ng of PCDNA 3.1+ empty vector with 1.5µl/well of Lipofectamine 2000 following manufacturer’s instructions (Invitrogen). After 6h of incubation, the media was then replaced with fresh growth medium and cells were cultured overnight until cycloheximide treatment on the next day.

### shRNA knockdown

3×10^6^ 293FT cells were transfected with 0.8μg of VSV-G, 5.4μg psPAX2, and 6μg of pLKO.1-shRNA plasmid (Openbiosystems, Chicago, United States) using lipofectamine 2000 (Invitrogen) according to the manufacturer’s instructions. shRNA mature antisense sequences for TRAF1 and PKN1 are AACAATGTTCTCAAACACACG and TATCCGCTTCTCACACATCAG, respectively. 60h after transfection, the supernatant of the cultures was collected, filtered through a low protein binding 0.22μM filter, and added to target cells for 24h before selection with 4μg/ml puromycin (Bio Basic, Markham, Canada). For TRAF1 knockdown in RAJI cells, cells were then seeded in 96 wells at 1 cell per well, allowing cells to grow into clones from a single cell. Several clones were then tested for reduction in TRAF1 levels by Western Blotting and Flow cytometry, and those with the lowest TRAF1 levels were chosen for downstream assays. For PKN1 knockdown in RAJI and 293T cells, sufficient reduction in PKN1 protein level was observed in the pool knockdown by Western blotting so no cloning was necessary.

### Cycloheximide Inhibition and Western blot

Following overnight recovery from transfection, 293FT or RAJI cells were treated or not with cycloheximide (Sigma-Aldrich) at 3μg/ml as indicted in the figures. After treatment, the media was removed and cells were washed once with PBS (Sigma-Aldrich), then lysed for protein extraction. Where indicated, SMAC mimetic BV6 (Sigma-Aldrich, St. Louis, USA) was added at 5μM. Cells were lysed in 0.5% Nonidet P-40 (Millipore-Sigma, Oakville, Canada) with phosphatase and protease inhibitor mix (Roche, Basel, Switzerland). Total protein concentration was quantified by a colorimetric assay (Bio-Rad, Berkeley, United States), then subjected to SDS-PAGE (10% gel) and transferred to polyvinylidene difluoride membranes (semi-dry transfer; Bio-rad).After blocking, membranes were probed with antibodies specific for TRAF1 (clone 45D3, Cell Signaling, Danvers, United States), PKN1 (BD Biosciences, Franklin Lakes New Jersey, United States) and GAPDH (Thermo Fisher Scientific), followed by HRP-conjugated anti-rabbit or anti-mouse (Jackson Immunoresearch, Baltimore, United States), and signals were detected with a chemiluminescence substrate (GE Healthcare, Baie D’Urfe, Quebec, Canada) and visualized by autoradiography.

### Immunoprecipitation

6×10^6^ RAJI cells in 10cm dishes were treated with Cycloheximide (see above) with or without BV6 and lysed in 800 μl 1% NP-40 buffer including protease and phosphatase inhibitor cocktails. 20 μl prewashed magnetic Protein G Dynabeads (Thermofisher) were incubated with 0.5 μg antibody for CD40 (IP1; G28.5, purified from hybridoma cell line using Protein G sepharose, hybridoma was originally provided by Diane Hollenbaugh, then at Bristol-Myers Squibb; the antibody is now available at BioXCell) for 10min at room temperature, washed and then incubated with 100 μl of lysates overnight at 4 °C. Unbound proteins were saved for a subsequent immunoprecipitation (IP2) by incubating them with 20 μl prewashed magnetic Protein G Dynabeads (Thermofisher) for 10min at room temperature and 0.5 μg antibody for TRAF1 (clone 1F342; Serviceeinheit Monoklonale Antikörper; Institut für Molekulare Immunologie, Munich, Germany). After three washes, immunoprecipitated proteins from IP1 and IP2 were fractionated on a 10% SDS-PAGE and immunoblotted for TRAF1 (clone 45D3; Cell Signaling).

### Kinase screen

A screen of the 700 kinase Ontario Institute for Cancer Research (OICR) inhibitor library at 1μM concentration on PKN1 kinase activity *in vitro* was conducted by contract research organization Eurofins as described on their website (www.eurofins.com). Briefly, PKN1 is incubated with 8 mM MOPS pH 7.0, 0.2 mM EDTA, 250 μM KKLNRTLSFAEPG, 10 mM magnesium acetate and gamma-^33^P-ATP. The reaction is initiated by the addition of the Mg/ATP mix. After incubation for 40 minutes at room temperature, the reaction is stopped by the addition of phosphoric acid to a concentration of 0.5%. 10 ul of the reaction is then spotted onto a P30 filtermat and washed four times for 4 minutes in 0.425% phosphoric acid and once in methanol prior to drying and scintillation counting. The kinase inhibitor library (**Table S2**) is an expanded version of one we have used previously^28, 29^ and is comprised both of clinically investigated or marketed agents, as well as pre-clinical compounds identified from the patent and primary literature. Of the 700 kinases screened, 28 were selected for further dose titration (**Table S3**). Of the 28 screened *in vitro* by Eurofins, 9 were selected for further testing on RAJI cells (**Table S4**). Inhibitors were added in a final concentration of 0.1% DMSO in complete media and TRAF1 was measured by intracellular flow cytometry at 24hrs. Known targets of these drugs and IC50 for inhibition of PKN1 kinase activity *in vitro* are included in **Table S3 and S4**.

### Treatment of RAJI cells with inhibitors and flow cytometry analysis

shCTL, shTRAF1 and shPKN1 RAJI cell lines, were seeded at 2×10^5^ cells per well in a 48-well plate in complete media (RPMI 1640 with added 10% Fetal Calf Serum (FCS), 2-ME, glutamine, penicillin, streptomycin and non-essential amino acids). Cells were harvested for flow cytometric analysis after 24h of treatment with 0.1% DMSO control, PF-941222^30^, UNC-2025 (Selleckchem, Houston, USA), crenolanib (Selleckchem), ipatasertib (Selleckchem), tofacitinib (Selleckchem), AT7867 (Selleckchem), uprosertib (Selleckchem), XL-228 (Chemietek, Indianapolis, USA), or OTSSP167 (Selleckchem), at various concentrations. All RAJI cells were treated with inhibitors in a final concentration of 0.1% DMSO in complete media. For analysis by flow cytometry, Fc receptors were blocked with human Fc Block (eBioscience). For surface staining, cells were stained with eBioscience Fixable Viability Dye eFluor® 506 and then fixed for intracellular staining with Foxp3/Transcription Factor Staining Buffer Set (eBioscience). Purified TRAF1 antibody (clone 1F3, Serviceeinheit Monoklonale Antikörper; Institut für Molekulare Immunologie, Munich, Germany) was used prior to secondary staining with goat anti-rat PE (clone poly4054, Biolegend) for TRAF1.

### OP9 co-culture, CLL samples and treatment with inhibitors

OP9 cells were re-suspended at 10^5^ cells/mL in OP9 Media (α-MEM with added 20% FCS, glutamine, penicillin and streptomycin) and seeded at 5×10^4^ cells per well into a 24-well plate. CLL patient samples were thawed and re-suspended at 8×10^6^ cells/mL in high glucose complete media (complete media with added 2.5g/L total glucose), plated at 2×10^6^ cells per well on a 24-well plate containing confluent OP9 and rested overnight. Samples were collected for flow cytometric analysis after 24h of treatment with 0.1% DMSO control or OTSSP167 or XL-228 or Venetoclax (EnzoLife Sciences, Farmingate, NY, ordered through Cedarlane, Ontario, Canada) at various concentrations. All CLL cells were treated with inhibitors in a final concentration of 0.1% DMSO in high glucose complete media.

### Flow cytometry analysis of patient samples

#### Bcl-2 family members and TRAF1

Anti-human Fc Block was used to block Fc receptors. For surface staining, live cells were stained with Fixable Viability Dye eFluor® 506. CLL cells were surface stained with anti-CD19 BV605 (clone HIB19) from Biolegend (San Diego, CA) and CD5 PE-Cy7 (clone UCHT2) purchased from eBioscience (La Jolla, CA). For intracellular staining, cells were fixed and permeabilized using Foxp3/Transcription Factor Staining Buffer Set (eBioscience). Purified TRAF1 antibody, anti-human Bcl-2 BV421 (clone 100, Biolegend) and anti-human Mcl-1 Alexa Fluor 647 (clone D2W9E, New England Biolabs) were used prior to secondary staining with goat anti-rat PE for TRAF1.

#### Phosphomarkers

For phosphoflow, live cells were stained with Fixable Viability Dye eFluor® 506 for 10min at 37°C prior to adding one volume of 2X Cytofix/Perm/Wash Buffer (3% PFA + 2X BD Perm/Wash + ddH20) directly to cell culture and incubated at room temperature for 15min and on ice for 1h. Cells were washed twice with Perm/Wash buffer (BD Biosciences) and re-suspended in Perm/Wash buffer with anti-human Fc blocking antibody. Cells were then stained with anti-CD19 BV605, anti-CD5 PE-Cy7, anti-human pNFκBp65-eFluor 660 (clone B33B4WP), anti-pS6 PE (clone cupk43k) and anti-pERK PerCP-eF710 (clone MILAN8R) from eBioscience for 1h at room temperature.

#### Cleaved caspase-3 and TRAF1

Anti-human Fc Block was used to block Fc receptors and then surface stained with Fixable Viability Dye eFluor® 506, anti-CD19 BV605 and anti-CD5 PE-Cy7. For intracellular staining, cells were fixed and permeabilized using the Foxp3/Transcription Factor Staining Buffer Set. Purified TRAF1 antibody and anti-cleaved caspase-3 Alexa Fluor 647 (clone D3E9, CST) were used prior to secondary staining with goat anti-rat PE (clone poly4054, Biolegend) for TRAF1. Gating on live cells was excluded when analysing cleaved caspase-3^+^ cells.

Data were acquired on a BD LSRFortessa™ X-20 flow cytometer and analyzed using FlowJo v10.7.1 software. Protein expression by flow cytometry is quantified as the median fluorescence intensity (MFI) of the specific antibody stain minus the fluorescence minus one (FMO; background) control, which is referred to as “dMFI”. Statistics were calculated using Prism GraphPad v8 software. Researchers were blind to patient demographics and clinical data until experimental data acquisition and analysis was complete.

### Data availability statement

Complete gels and raw flow cytometry FCS files associated with the figures are available from the authors upon request.

## Results

### PKN1 is required for TRAF1 protein stability

To determine the effect of PKN1 on TRAF1 biology, we knocked down PKN1 in lymphoma cell lines using lentiviral delivery of either a control small hairpin (sh) RNA (shCTL) or an shRNA targeting PKN1 (shPKN1). Unexpectedly, knockdown of PKN1 led to a concomitant reduction in the level of TRAF1 protein in RAJI and Daudi cells (**Figure 1A**). To test whether PKN1 affected protein stability, RAJI cells expressing control shRNA (shCTL) or shRNA targeting PKN1 (shPKN1) were incubated with cycloheximide at 37°C to block new protein synthesis and analyzed for levels of PKN1 and TRAF1 by western blotting. The results show that the absence of PKN1 is associated with decreased stability of TRAF1 protein (**Figure 1B**).

**Figure 1.**
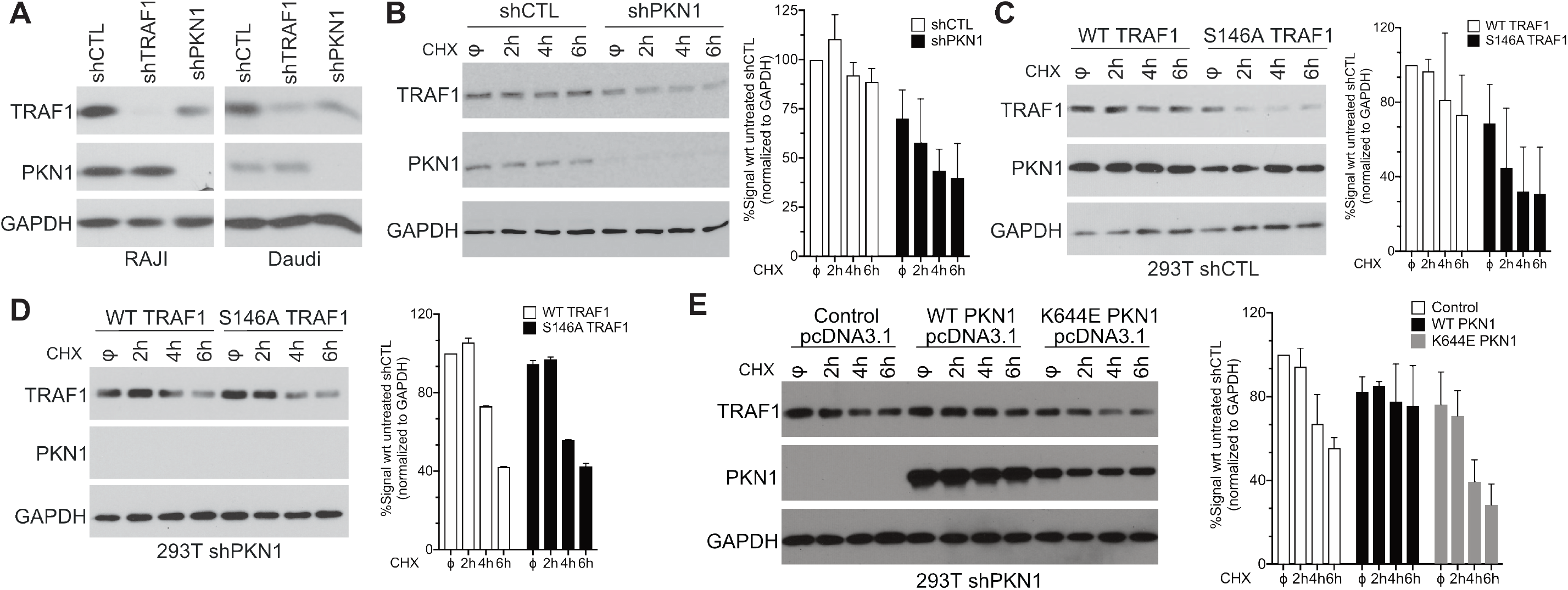
PKN1 kinase activity and TRAF1 S146 are required for TRAF1 protein stability. (A) RAJI or Daudi cells were stably transduced with lentiviruses expressing control shRNA (shCTL), with shRNA targeting TRAF1 (shTRAF1) or with shRNA targeting PKN1 (shPKN1) and whole cell lysates were subjected to western blot analysis for TRAF1, PKN1 or GAPDH. (B) shCTL or shPKN1 RAJI cells were treated with cycloheximide for the indicated times in hours (h), then whole cell lysates were subjected to western blot to determine TRAF1, PKN1 or GAPDH levels as indicated on the figures (C,D). 293T cells stably transduced with shCTL (C) or shPKN1 (D) lentiviruses were transiently transfected with WT TRAF1 or with TRAF1 S146A expression plasmids. Cells were treated or not with cycloheximide (CHX) for the indicated times, then whole cell lysates were subjected to Western blot analysis of TRAF1, PKN1 or GAPDH levels. (E) shPKN1 293T cells were transiently co-transfected with TRAF1 as well as shPKN1-resistant WT or K644E (kinase dead) PKN1. Cells were then treated with cycloheximide (CHX) and then cell lysates were subjected to Western blot to analyze TRAF1, PKN1 or GAPDH levels. All experiments were repeated at least two times. Densitometry results are indicated to the right of each gel.

PKN1 was previously shown to phosphorylate TRAF1 on serine 146.^27^ To determine if the phosphorylation of TRAF1 S146 by PKN1 is responsible for TRAF1 stability, we generated a mutant human TRAF1 construct in which serine 146 was replaced with alanine (S146A). 293 cells were chosen for this experiment as they do not express TRAF1.^31^ Upon transient transfection into 293T cells, TRAF1 S146A showed reduced stability compared to WT TRAF1 (**Figure 1C**). In contrast, when TRAF1 WT or S146A were expressed in 293T cells in which PKN1 had been knocked down by shRNA, both WT and TRAF1S146A showed a similar half-life, which was decreased compared to that of WT TRAF1 in PKN1 sufficient cells (**Figure 1D**).

To test whether the kinase activity of PKN1 was required for TRAF1 stability, we expressed TRAF1 as well as shRNA-resistant WT or kinase dead (K644E) PKN1^32^ in 293T cells that had been stably transduced with shPKN1 (**Figure 1E**). In the absence of PKN1, TRAF1 protein showed a half-life of between 2 and 4h. Overexpression of WT PKN1 in the PKN1 knockdown cells increased the TRAF1 protein half-life to at least 6h. In contrast, kinase dead PKN1 failed to increase TRAF1 stability. Although we observed higher expression of WT compared to kinase dead PKN1 in 293T cells, the level expressed was still in excess of the physiological level of PKN1 observed in cells before knockdown. Moreover, in another experiment, supraphysiological levels of kinase dead PKN1 also failed to increase TRAF1 stability (data not shown). Thus, reduced expression of PKN1 K644E is unlikely to account for its lack of efficacy in conferring TRAF1 protein stability. The finding that WT but not K644E PKN1 restores TRAF1 stability argues against off -target effects of PKN1 knockdown. Taken together, these results show that phosphorylation of TRAF1 by PKN1 on S146 is required for TRAF1 protein stability in a B cell lymphoma.

### PKN1 protects TRAF1 from cIAP-mediated degradation in the CD40 signaling complex

TRAF1 is important for the recruitment of cIAPs leading to NF-κB activation. However, cIAPs can add K48-linked ubiquitin to TRAF proteins, thereby inducing TRAF protein degradation.^20^ Therefore, we hypothesized that PKN1 is required to protect TRAF1 from cIAP mediated degradation during CD40 signaling. SMAC mimetics inhibit cIAP function by inducing their autoubiquitination and degradation.^33^ To test the role of cIAPs in TRAF1 degradation, RAJI cells were treated with cycloheximide with or without the SMAC mimetic BV6 and then CD40 was immunoprecipitated from control RAJI cells or from RAJI cells knocked down for PKN1. A second immunoprecipitation of TRAF1 was used to analyze the effect of SMAC mimetics on the cytosolic, non-CD40 associated TRAF1 pool. In the presence of PKN1, CD40 associated TRAF1 protein was decreased at 6h, but rescued by BV6 (**Figure 2A**). In the absence of PKN1, TRAF1 was less stable, already degrading by 3h, and again partially rescued by BV6 treatment. In contrast, the cytosolic pool of TRAF1 (IP2 in **Figure 2A**) was not sensitive to BV6 treatment. These results show that cIAPs contribute to degradation of TRAF1 in the CD40 signaling complex, and that PKN1 partially protects TRAF1 from cIAP-induced degradation.

**Figure 2.**
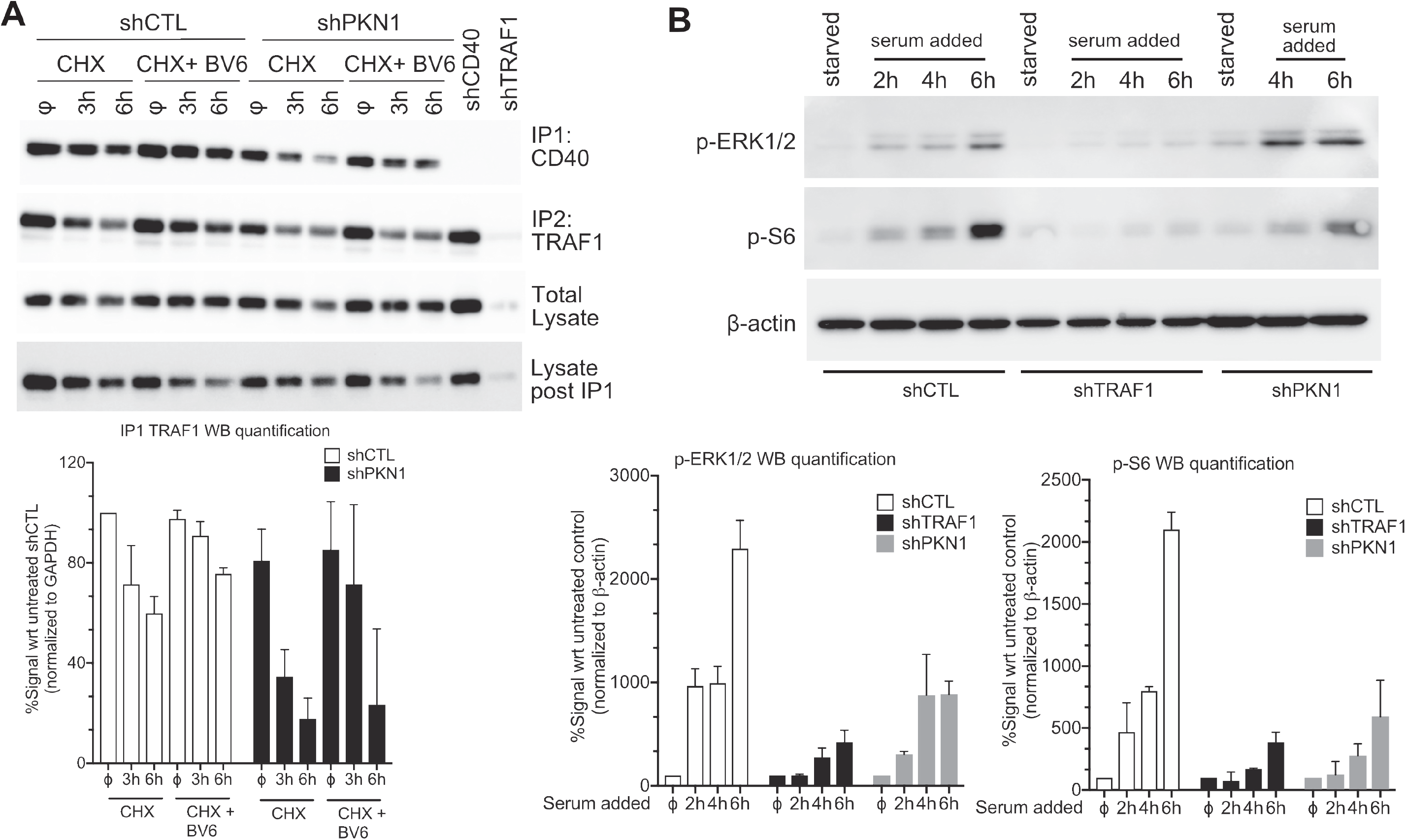
PKN1 is required to protect TRAF1 from cIAP-mediated degradation during CD40 signaling in RAJI cells, thereby contributing to TRAF1 dependent signaling. (A) shCTL or shPKN1 RAJI cells were treated with cycloheximide (CHX) for the indicated times, with or without SMAC mimetic BV6 treatment, then the CD40 signaling complex was immunoprecipitated from whole cell lysates (IP1). TRAF1 was then immunoprecipitated from the supernatants of the CD40 immunoprecipitates (IP2). IP1, IP2 or whole cell lysates were analyzed for levels of TRAF1 by Western blotting. (B) shCTL, shTRAF1 or shPKN1 RAJI cells were serum starved for 24h to reduce constitutive signaling then serum was added back for the indicated times and whole cell lysates were analyzed by Western blotting for levels of p-ERK1/2, p-S6 or β-actin as indicated in the figure. Each panel has been repeated at least 3 times. Densitometry results indicated below the figure and are reported as the average of two similar experiments.

### TRAF1 and PKN1 contribute to constitutive MAPK and mTOR signaling in RAJI cells

We next asked whether PKN1 and TRAF1 contribute to constitutive signaling in RAJI cells. RAJI cells were deprived of serum and amino acids and then serum / amino acids were added back to synchronize constitutive signaling. Knockdown of TRAF1 reduced the level of pErk1/2 and pS6, a downstream target of mTOR (**Figure 2B**). Knockdown of PKN1 had an intermediate effect compared to direct TRAF1 knockdown, correlating with its partial effect on TRAF1 levels (see **Figure 1A**). These findings suggested that inhibition of PKN1 could be used to reduce constitutive TRAF1-dependent survival signaling in B cell cancers.

### Identification of PKN1 inhibitors that reduce TRAF1 among the OICR library of 700 kinase inhibitors

The above findings showed that PKN1 is important for TRAF1 protein stability. Therefore, it was of interest to identify a PKN1 inhibitor that could be used to test the importance of PKN1 in primary unmanipulated patient samples. To this end, we conducted a screen of the Ontario Institute for Cancer Research (OICR) kinase inhibitor library, which consists of 700 known kinase inhibitors. After an initial screen of inhibitors at 1μM (**Table S2**), 28 kinase inhibitors that substantially inhibited PKN1 were subjected to further dose-response analysis (**Table S3**). Of these compounds, we chose 9 that inhibited PKN1 *in vitro* in the 11-50nM range to test for effects on lowering TRAF1 in RAJI cells. The other known targets and IC50 for PKN1 inhibition for these compounds are listed in **Table S4**. The choice of compounds for testing on RAJI was made to avoid compounds for which known off target effects make them likely to decrease TRAF1 through inhibition of NF-κB. A further selection was made based on their inherent kinase selectivity or the uniqueness of the core scaffold for future drug development considerations. To validate the measurement of TRAF1 by flow cytometry, we used RAJI shCTL, shTRAF1 and shPKN1 cell lines (**Figure 3A**). Control RAJI cells showed a high level of TRAF1 by flow cytometry, shTRAF1 cells showed minimal TRAF1 expression and shPKN1 RAJI cells showed an intermediate level of TRAF1, consistent with the Western blot data in Figure 1. Six of the nine compounds tested, PF-941222 (IC50 for reducing TRAF1= 4.0μM), ipatasertib (IC50= 69μM), AT7867 (IC50= 1.4μM), uprosertib (IC50= 4.9μM), XL-228 (IC50= 2.3μM) and OTSSP167 (IC50= 18nM), induced dose-dependent decreases in TRAF1 protein after 24h of treatment of RAJI cells (**Figure 3B**). Two of the compounds, UNC-2025 and crenolanib increased rather than decreased TRAF1 levels, and tofacitinib, a JAK inhibitor previously shown to inhibit PKN1^34^, had minimal effects on TRAF1, potentially due to effects on other kinases. OTSST167, previously known as a MELK inhibitor, was the most potent of the compounds in causing dose-dependent loss of TRAF1. At the highest doses of OTSST167, when TRAF1 levels went below 10% of the original level, we saw induction of cell death in RAJI cells, as evidenced by activated caspase 3 (**Figure 3C**), albeit about half the cells were resistant to cell death. We next chose OTSST167, as well as a second PKN1 inhibitor that lowered TRAF1, XL-228, for effects on constitutive signaling in RAJI cells (**Figure 3D**). The loss of TRAF1 in RAJI cells induced by the two inhibitors was associated with dose-dependent loss of pS6 and pNF-κB p65. pERK levels were decreased only at the highest levels of OTSSP167 but not by XL-228. Taken together these data show that inhibitors selected for their ability to inhibit PKN1 and reduce TRAF1 in RAJI cells also reduced constitutive signaling in RAJI, with similar effects as knockdown of PKN1 or TRAF1.

**Figure 3.**
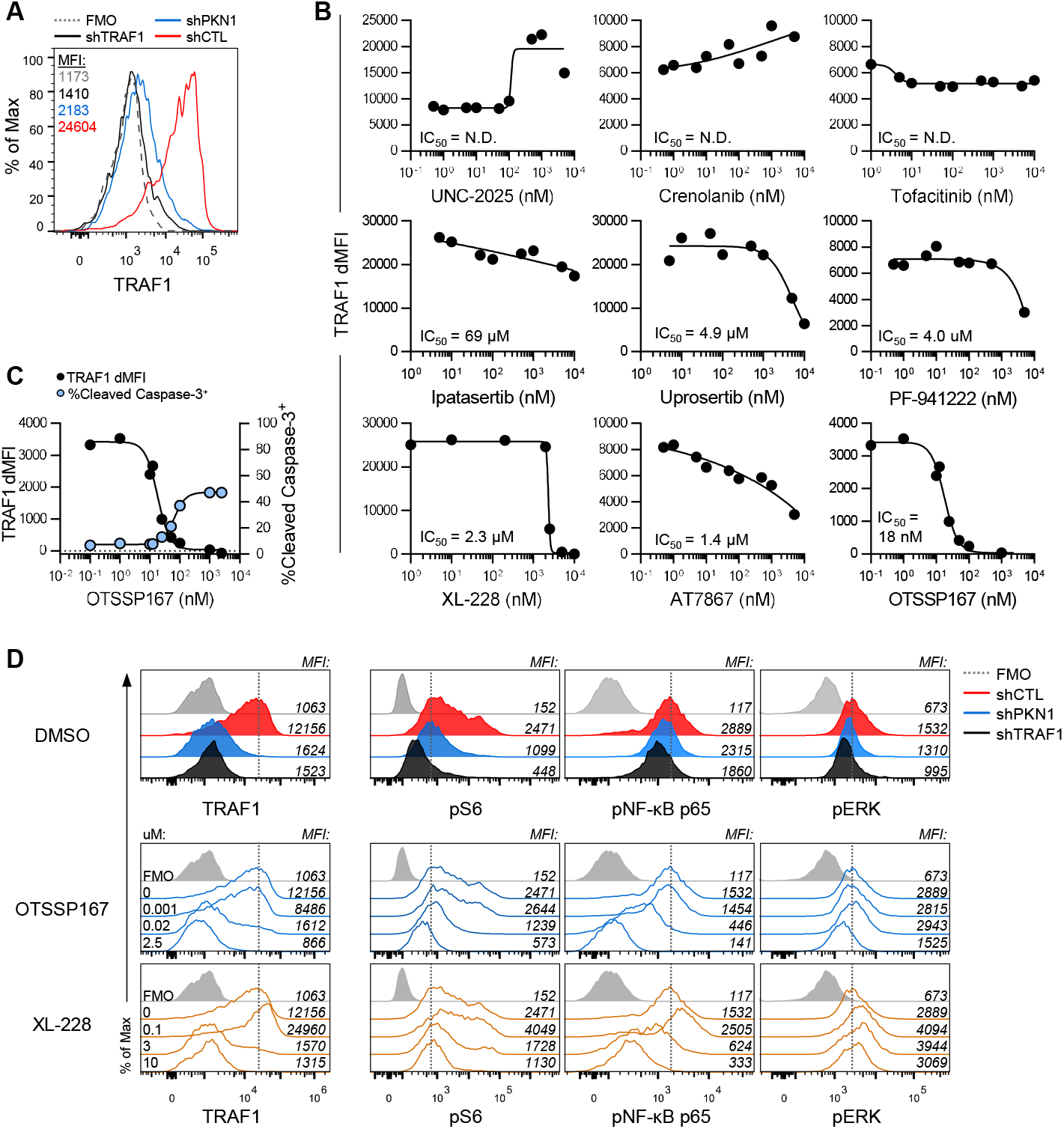
Identification of PKN1 inhibitors that decrease TRAF1 protein levels and constitutive signaling in RAJI cell lines. (A) Base line expression of TRAF1 in shCTL, shTRAF1 and shPKN1 RAJI cell lines, representative of 7 independent experiments. (B) shCTL RAJI cells were treated with the indicated inhibitor at doses ranging from 0.1nM-10μM for 24h. TRAF1 expression was measured by intracellular flow cytometry after treatment. dMFI refers to the MFI for the TRAF1 stain minus the FMO (background) control. To calculate the IC50 for PF-941222, AT7867, ipatasertib and uprosertib where TRAF1 expression was not maximally reduced, TRAF1 was assumed to reach 0 dMFI. N.D. = not determined. (C). Effect of OTSST167 on TRAF1 levels and death of RAJI cells measured by activated caspase 3. (D) Baseline expression of TRAF1 and phosphorylation of signaling intermediates pS6, pNF-κBp65 and pERK in shCTL, shPKN1 and shTRAF1 RAJI cell lines (top). shCTL RAJI cells were treated with OTSSP167 or XL-228 at the indicated concentration for 24h. TRAF1, pS6, pNF-κBp65 and pERK were measured by flow cytometry (middle, bottom). Data are representative of 2 independent experiments.

### PKN1 is constitutively phosphorylated on Thr744 in primary CLL cells

To examine PKN1 expression in primary CLL, we obtained frozen PBMC from CLL patients from the Leukemia Tissue Bank of Princess Margaret Hospital /UHN (**Table S1**). For 8/8 patient CLL samples examined, there was constitutive, relatively uniform expression of PKN1 (**Figure S1A and S1B**). PKN1 can be activated by phosphorylation of its activation loop by the kinase PDK1.^35, 36^ Of interest, this activated pThr774 form of PKN1 was detected in all 8 CLL patient samples examined (**Figure S1A and S1B**), whereas there was lower expression of pThr774 PKN1 in purified B cells from healthy donors (**Figure S1B**). Phospho-Thr 774 is also observed in RAJI cells (data not shown). All CLL samples examined also express TRAF1 protein as well as Zap70, albeit with some variation between samples. These data suggest that at least a fraction of PKN1 is found in its active, phospho-Thr744 state in CLL cells with minimal pThr744 PKN1 detected in healthy donor B cells, consistent with a possible role for PKN1 in contributing to TRAF1 overexpression in CLL.

### OTSSP167 and XL-228 reduce TRAF1 and increase cell death in most primary CLL samples tested

Elevated expression of TRAF1 in hematopoietic malignancies,^21^ combined with evidence in the literature that TRAF1 protects lymphocytes from apoptosis and contributes to lymphomagenesis in mice,^26, 37-39^ suggests that TRAF1 may contribute to apoptosis-resistance in CLL. However, our attempts to knockdown TRAF1 in primary CLL with the shRNA-lentiviral construct used to knockdown TRAF1 in RAJI cells was unsuccessful, as no viable cells were recovered with lowered TRAF1, potentially due to the importance of TRAF1 in CLL survival. Therefore, we chose 3 PKN1 inhibitors, OTSSP167, XL-228 and AT7867, identified on the basis of their effect on PKN1 in vitro, and their ability to decrease TRAF1 in RAJI cells for their effects on primary CLL patient cells. We monitored intracellular TRAF1 by flow cytometry 24h after treatment and activated caspase-3, as an indicator of apoptotic cell death. CLL cells in the PBMC cultures were identified by co-expression of CD5 and CD19 (**Figure 4A**). TRAF1 expression was determined by gating on live CD5^+^CD19^+^ CLL cells, while frequency of apoptotic cells was determined by the gating on cleaved caspase-3^+^ cells from CD5^+^CD19^+^ single cells without the live gate. OTSSP167 and XL-228 each induced dose-dependent decreases in intracellular TRAF1 protein and increases in activated, cleaved caspase-3 for 16/19 and 9/10 CLL patient samples analyzed, respectively (**Figure 4B-C and data not shown**). Moreover, the IC50 for decreasing TRAF1 protein levels was similar to the IC50 for activating caspase-3 for both OTSSP167 (30.7nM and 33.9nM, respectively) and XL-228 (2.96μM and 2.54μM, respectively) (**Figure 4B-C**), consistent with the linkage of the two effects. Two outliers (**Figure S2**) had low TRAF1 to start with and showed a slight increase in TRAF1 with treatment. A third outlier gave equivocal results, with decreases in TRAF1 only at high doses of OTSSP167 (**Figure S2**). Treatment with the lower affinity inhibitor AT7867 had no effect on TRAF1 expression or cell death in any of the samples (**Figure S3A)**. Analysis of CLL cells at suboptimal concentrations of OTSSP167 (50nM) and XL-228 (4μM), where approximately 20-50% of cells survived treatment, showed a clear dichotomy of cells into a TRAF1^lo^ and TRAF1^hi^ subset. Consistent with a role for TRAF1 in CLL survival, activated caspase-3 was limited to the TRAF1^lo^ subset of cells within each donor (**Figure 4 D-E**). Together, these results show that for the majority of primary patient CLL samples tested, treatment with OTSSP167 and XL-228 leads to a reduction in TRAF1 protein in CLL cells. The loss of TRAF1 was not secondary to cell death, as TRAF1 was measured in the live cell fraction only. Thus, loss of TRAF1 correlates with increased CLL death.

**Figure 4.**
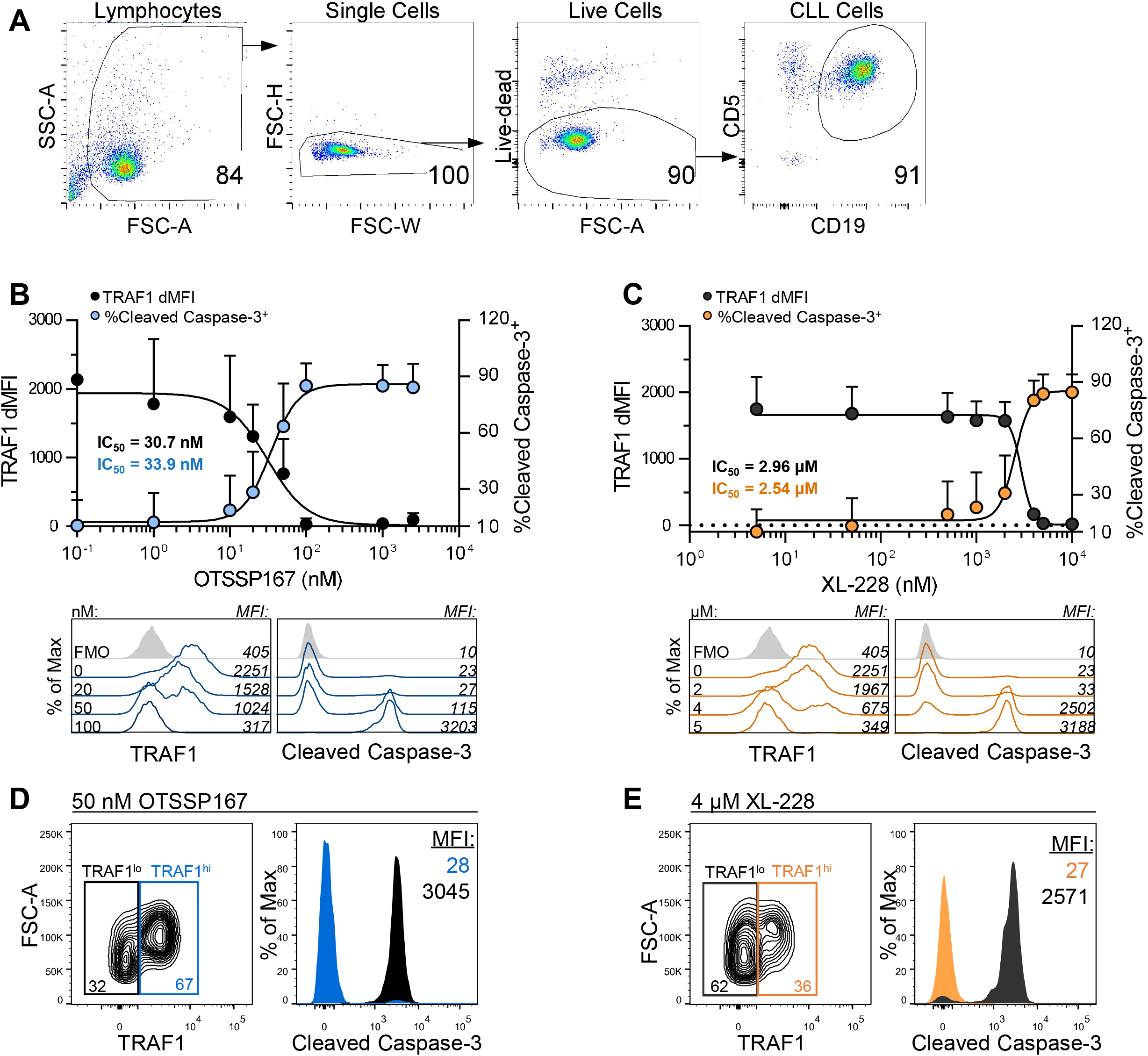
OTSSP167 and XL-228 decrease TRAF1 protein levels and increase cell death in primary CLL cells. (A) Representative gating strategy to identify CLL cells. TRAF1 and cleaved caspase-3 expression were measured in CLL cells by flow cytometry after treatment with DMSO control or (B) OTSSP167 or (C) XL-228 for 24h at the indicated concentrations. In panels (B) and (C), line graphs (top) show TRAF1 dMFI and %cleaved caspase-3^+^ cells as mean ± SD. Histograms from one representative donor depict TRAF1 MFI (bottom left) and cleaved caspase-3 MFI (bottom right) in CLL cells after treatment with DMSO or inhibitors. TRAF1 MFI was determined by gating on Live CD5+CD19+ CLL cell. Frequency of cleaved caspase-3+ cells was determined by gating on CD5+CD19+ single cells. (D, E) (B, D) n=6 CLL patient samples, 2 experiments pooled. Representative of a total of 16 donors across 8 total experiments. (C, E) n=6 CLL patient samples, 2 experiments pooled. Representative of a total of 9 donors across 4 total experiments.

### OTSSP167 and XL-228 decrease the expression of survival molecules in primary CLL cells

TNFR family members can upregulate pro-survival Bcl-2 family members via NF-κB signaling.^40^ Consistently, 24h after the addition of OTSSP167 and XL-228, Bcl-2 and Mcl-1 protein levels were reduced in CLL cells in a dose-dependent manner (**Figure 5A-B**). Furthermore, the IC50s for lowering Bcl-2 and Mcl-1 by OTSSP167 (49.8nM and 25.7nM, respectively) and XL-228 (3.52μM and 2.54μM, respectively) were similar to the IC50s for decreasing TRAF1 and increasing caspase-3 activation (**Figure 4B-C and Figure 5A-B**). We also examined Bcl-XL, but the flow cytometry signal in untreated samples was marginal and therefore not analyzed further (data not shown). AT7867, which did not affect TRAF1 and cell death in CLL cells also did not affect Bcl-2 and Mcl-1 expression (**Figure S3B**). These results show that the kinase inhibitors OTSSP167 and XL-228 reduce the level of TRAF1 protein in primary CLL cells and this decrease is associated with concomitant reductions of Bcl-2 and Mcl-1.

**Figure 5.**
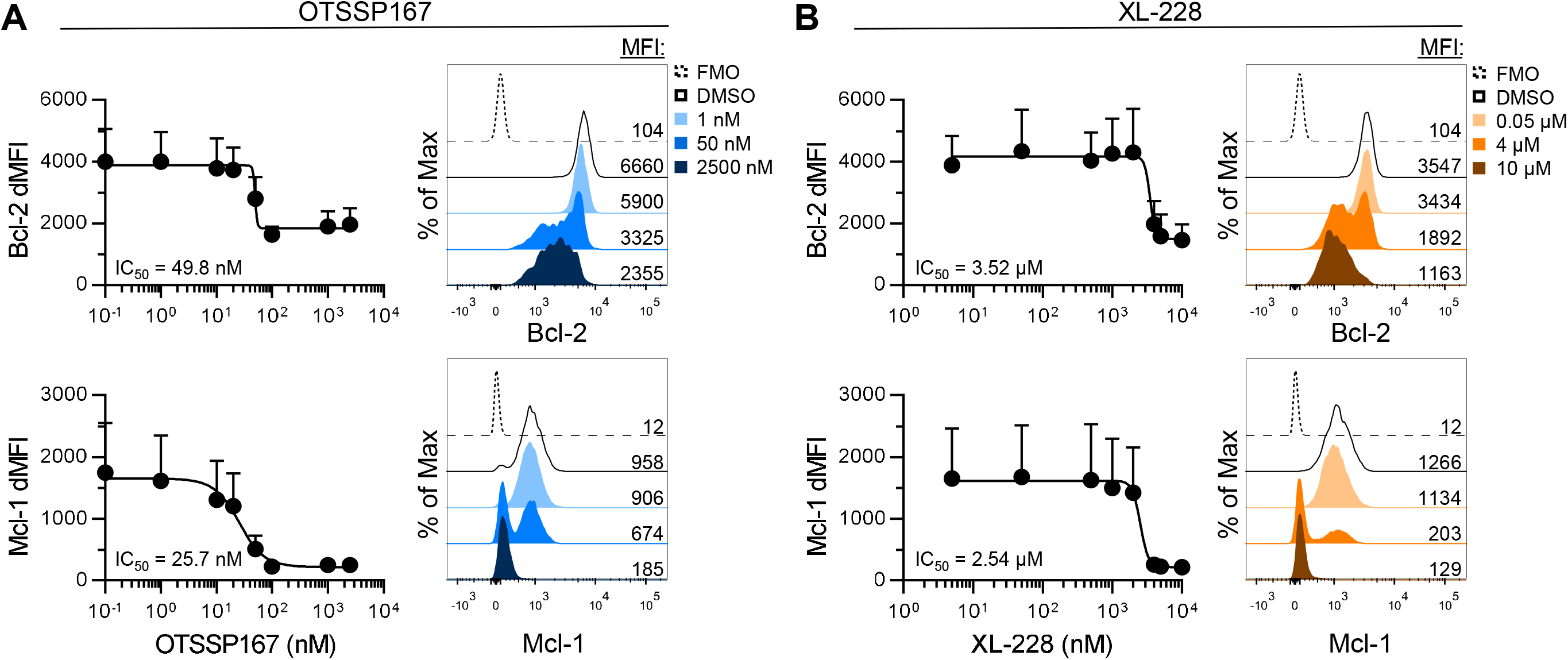
OTSSP167 and XL-228 decrease the level of Bcl-2 and Mcl-1 protein expression in primary CLL cells. Bcl-2 and Mcl-1 expression were measured in live CLL cells after treatment with DMSO control or (A) OTSSP167 or (B) XL-228 for 24h at the indicated concentrations. In each panel, line graphs (left) show mean dMFI ± SD, n=6 CLL patient samples (2 independent experiments pooled), and histograms (right) from a representative donor depict the MFI of Bcl-2 or Mcl-1 in CLL cells after treatment with DMSO or the indicated concentration of inhibitor. The same 6 CLL samples are shown here as in figure 4.

### OTSSP167 and XL-228 decrease signaling intermediates in primary CLL cells

TRAF1 enhances NF-κB, MAPK and pS6 activation downstream of TNFRs such as CD40, CD30, TNFR2, 4-1BB and the EBV encoded TNFR LMP1.^16, 39, 41-46^ Therefore, we asked how reduction of TRAF1 impacted these downstream signaling pathways in live, primary CLL cells. After treatment of CLL cells with OTSSP167 for 24h, we observed a dose-dependent decrease in pNF-κB p65 (IC50 = 0.32μM), pS6 (IC50 = 0.31μM) and pERK (IC50 = 1.04μM) (**Figure 6A**). Likewise, treatment of CLL cells with XL-228 resulted in a dose-dependent decrease in pNF-κB (IC50 = 2.47μM), pS6 (IC50 = 1.44μM) and pERK (IC50 = 2.23μM) (**Figure 6B**). In contrast, treatment of cells with AT7867 did not affect levels of pNF-κB, but slightly reduced pS6 and pERK (**Figure S3C**). As AT7867 had no effect on decreasing TRAF1 in primary CLL cells, the lack of effect on pNF-κB p65 is expected. As summarized in Table S4, AT7867 also inhibits AKT1,2 and 3 as well as S6K, possibly explaining the effects on pS6. These results show that the decreases in TRAF1 observed in primary CLL cells after treatment with PKN1 inhibitors are associated with a reduction in levels of pNF-κB p65, pS6 and pERK and largely recapitulate the results seen in RAJI cells.

**Figure 6.**
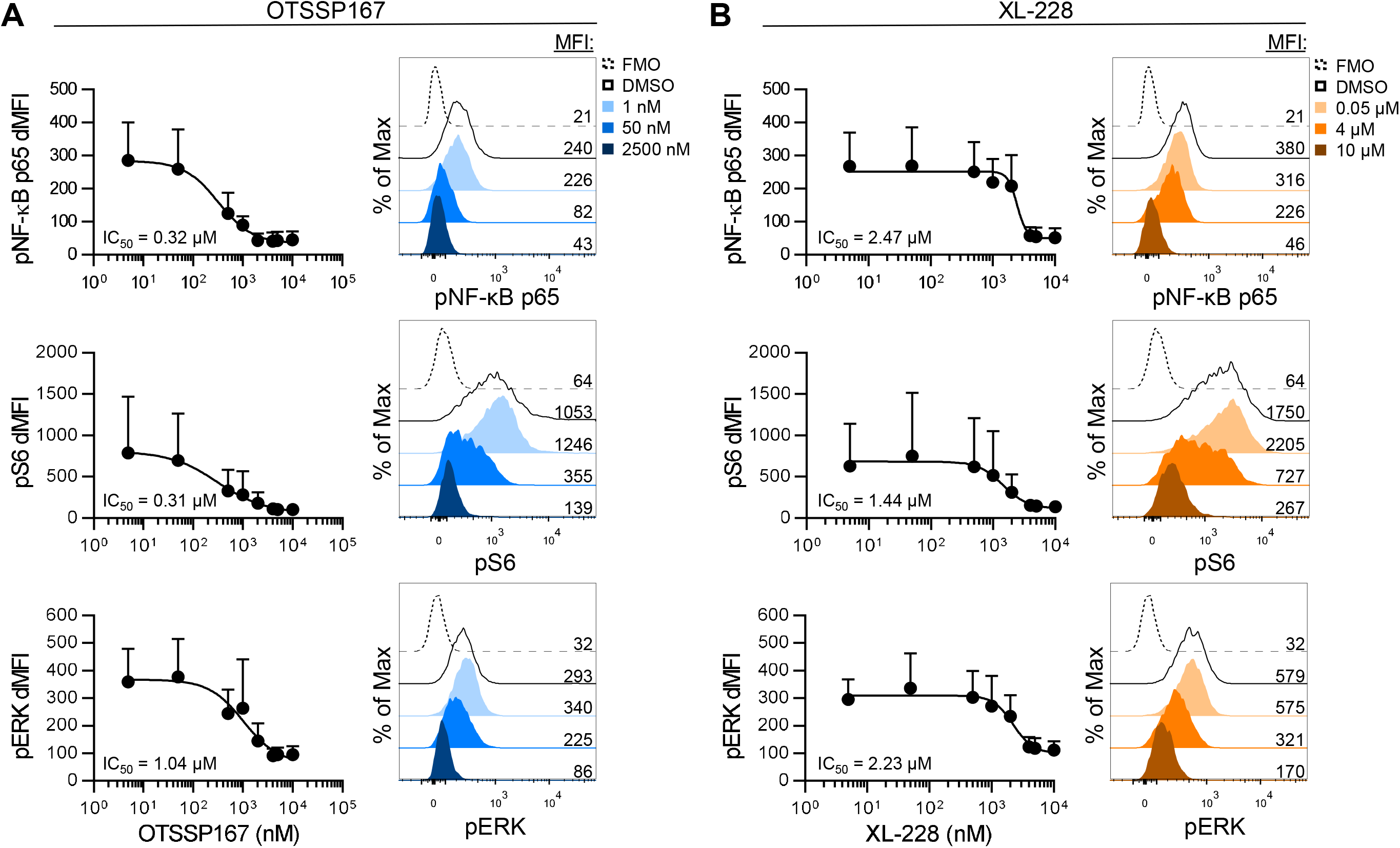
OTSSP167 and XL-228 decrease the level of signaling intermediates in primary CLL cells. Phosphorylated signaling intermediates pS6, pNF-κBp65 and pERK were measured in live CLL cells after treatment with DMSO control or (A) OTSSP167 or (B) XL-228 for 24h at the indicated concentrations. In each panel, line graphs (left) show mean dMFI ± SD, n=6 CLL patient samples (2 independent experiments pooled), and histograms (right) from a representative donor depict the MFI of pS6, pNF-κBp65 or pERK in CLL cells after treatment with DMSO or the indicated concentration of inhibitor. The same 6 CLL samples were used here as in figures 4 and 5.

### PKN1 inhibition in combination with venetoclax results in increased CLL death correlating with reduced levels of TRAF1 and Bcl-2 family proteins

Venetoclax, a BH3 mimetic that antagonizes Bcl-2 and induces apoptosis is approved for the treatment of patients with relapsed and 17p deleted CLL.^7, 47^ Although showing great promise, mechanisms of venetoclax resistance are emerging.^48, 49^ The finding that inhibitors that targeted TRAF1 also reduced Bcl-2 and Mcl-1 levels in CLL cells suggested these inhibitors might be complementary to venetoclax. Therefore, we examined effects of the stronger of the two inhibitors, OTSSP167, in combination with venetoclax. Fewer than 15% of primary CLL cells died when treated with suboptimal OTSSP167 (10nM) or venetoclax (1nM) alone (**Figure 7A**). However, when cells were treated with increasing concentrations of venetoclax in combination with suboptimal (10nM) OTSSP167, the IC50 for cell death (1.71nM) was decreased 5.8-fold compared to venetoclax treatment alone (9.91nM) (**Figure 7B**). However, when cells were treated with increasing amounts of OTSSP167 in combination with suboptimal (1nM) venetoclax, the IC50 for cell death was only reduced 1.9-fold to 23.7nM from 45.9nM with OTSSP167 treatment alone (**Figure 7C**). At the dose of venetoclax that resulted in approximately 50% death, we also observed that CLL cells from 4 of 6 patients were enriched for TRAF1^hi^ cells after venetoclax treatment. The addition of suboptimal OTSSP167 in combination with this dose of venetoclax eliminated this TRAF1^hi^ subset (**Figure 7D**). Further analyses of the TRAF1^hi^ and TRAF1^lo^ subsets showed that TRAF1^hi^ cells have higher expression of Bcl-2 family members Mcl-1 and Bcl-2 than TRAF1^lo^ cells (**Figure 7E**). Given the lack of effect of venetoclax in lowering TRAF1, this may explain the reduced effects on cell death when suboptimal venetoclax is added to OTSSP167 as OTSSP167 alone effectively eliminates both TRAF1^lo^ and TRAF1^hi^ populations (**Figure 7 C, F**). Taken together, these data show that by targeting TRAF1, the inhibitor OTSSP167 has complementary effects with venetoclax on CLL cells by eliminating TRAF1^hi^, Bcl-2 and Mcl-1 co-expressing venetoclax-resistant cells (**Figure 7G**). Moreover, measurement of TRAF1 provides a useful marker of the effectiveness of OTSSP167 in inducing death CLL cells.

**Figure 7.**
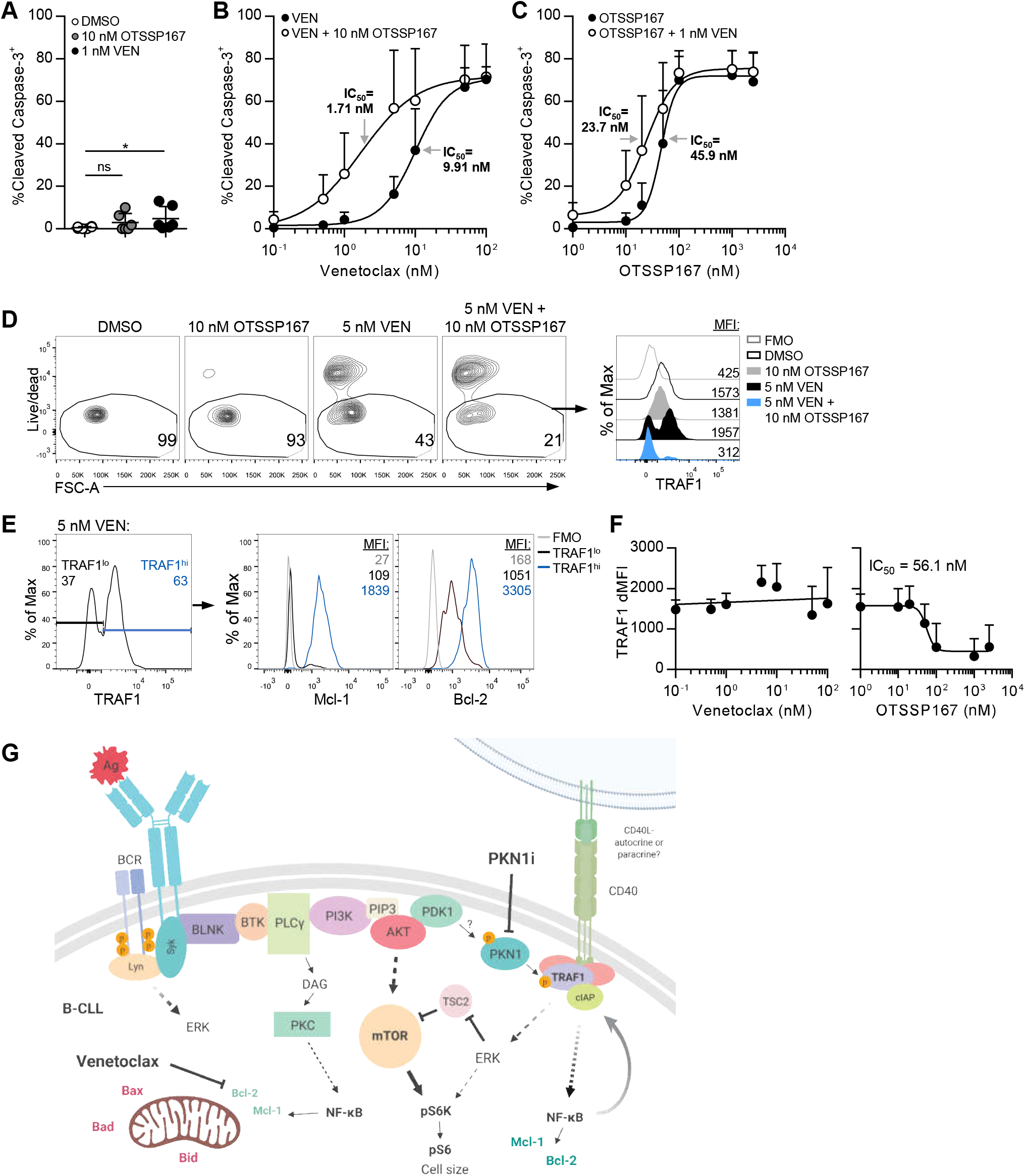
Increased cell death following combined treatment with OTSSP167 and venetoclax. Primary CLL cells were cultured on OP9 stromal cells and cleaved caspase-3 expression was measured by flow cytometry following 24h of treatment with: (A) DMSO, 10 nM OTSSP167 or 1 nM venetoclax, (B) 0.1 nM-100 nM VEN ± 10 nM OTSSP167 or (C) 1 nM-2.5 μM OTSSP167 ± 1 nM VEN. (D) Representative plots from one donor showing gating for live CLL cells and TRAF1 expression by live CLL cells. (E) Live CLL cells were gated on TRAF1^lo^ and TRAF1^hi^ cells for analysis of Bcl-2 and Mcl-1 expression. (F) TRAF1 expression on live CLL cells after treatment with the indicated dose of ventoclax (left) or OTSSP167 (right). Graphs show %cleaved caspase-3^+^ cells as mean±SD, n=6 (2 independent experiments pooled). 1 CLL sample was from figure 4-6, 5 were new samples. Statistical analysis was performed using a non-parametric Dunn’s multiple comparisons test. *p<0.05; ns, not significant. (G) Schematic showing BCR and TNFR signaling pathways in CLL, with sites of action of PKN1 inhibition and venetoclax indicated.

## Discussion

PKN1, a ubiquitous kinase, has been implicated as an effector of Rho GTPases ^50, 51^ and in regulation of cell migration.^52-55^ Here, we reveal another role for PKN1 in maintaining the stability of TRAF1 protein in lymphoma and CLL cells. Knockdown and restoration experiments in RAJI and 293 cells confirmed the need for PKN1 phosphorylation of TRAF1 S146 to prevent TRAF1 degradation during constitutive CD40 signaling in RAJI cells. During TNFR family signaling, TRAF1 is important in the recruitment of cIAPs for downstream NF-κB and MAPK signaling.^39^ As cIAPs can add K48-linked polyubiquitin to trigger protein degradation as well as K63-linked polyubiquitin to scaffold downstream signaling, we hypothesized that phosphorylation of TRAF1 by PKN1 is required to protect it from cIAP-induced degradation during constitutive TNFR family signaling in B cell related cancer cells. The finding that SMAC mimetics that induce degradation of cIAPs abrogated TRAF1 degradation in the CD40 signalosome with little impact on the cytosolic TRAF1 pool supports this hypothesis. PKN1 is regulated by phosphorylation on its activation loop by PDK1.^35, 36^ This active pThr774 form of PKN1 was readily detected by western blot in CLL patient samples, whereas substantially lower levels were detected in normal B cells from healthy donors. As PDK1 can be activated downstream of BCR signaling, the activation of PKN1 may be explained by constitutive BCR signaling in CLL cells.^56^ Thus, the presence of TRAF1 and the pThr744 form of PKN1 might be a useful adjunct in CLL screening. Interestingly, PDK1 and active PKN1 have also been noted in other cancers, with particularly high expression in ovarian serous carcinoma, suggesting that inhibition of PKN1 might have broader use.^57^

TRAF1 is an NF-κB induced protein that enhances NF-κB signaling downstream of TNFRs. Here we showed that decreasing TRAF1 by inhibition of PKN1 has the potential to break this feed-forward NF-κB signaling loop in TNFRSF-dependent cancers. Our study identified two compounds, OTSSP167 and XL-228, that when added to primary patient CLL cells induced dose dependent loss of TRAF1, pS6, pNF-κB p65, pERK, Mcl-1 and Bcl-2, and concomitant induction of cell death. This was not secondary to cell death, because we gated on live cells for all measurements except activated caspase 3 which was measured on the total population of CLL cells. Moreover, in RAJI cells, cell death was observed only when TRAF1 levels were reduced to minimal levels and RAJI death plateaued at 50%. This suggests RAJI may have compensatory mechanisms for survival compared to CLL, which show 100% loss of viability at high doses of the inhibitors. The correlation between decreased TRAF1 protein levels, decreased downstream signals and increased caspase-3 activation at similar IC50, is consistent with a direct effect through the pPKN1-pTRAF1-NF-κB/MAPK/pS6 signaling axis. In contrast, another inhibitor AT7867, with a higher IC50 for inhibiting PKN1 failed to decrease TRAF1 significantly in CLL cells and showed no effect on the downstream signals or cell death. OTSSP167 is also known as a MELK inhibitor. OTSSP167 has been tested in phase 1/2 trials for breast cancer and acute myeloid leukemia, and has been suggested as a CLL treatment based on its ability to induce apoptosis in primary CLL cells.^58^ However, many inhibitors, and in particular OTSSP167, have broad effects on multiple kinases.^59^ Thus, it is possible that the effects of OTSSP167 on PKN1 contributed to CLL cell death in MELK inhibitor studies.^58^ The finding that OTSSP167 and XL-228 show an order of potency for inhibiting TRAF1 and its downstream signals that matches the order of potency for PKN1 inhibition is also consistent with these drugs acting on PKN1. OTSSP167, with 11 nM IC50 for PKN1 inhibition *in vitro* and an IC50 for decreasing TRAF1 and increasing apoptosis of CLL cells of 30 nM, represents a good starting point for future drug discovery efforts to identify a more potent and selective PKN1 inhibitor. Other drugs identified as potential PKN1 inhibitors *in vitro*, such as the JAK inhibitor tofacitinib,^34^ were not effective in lowering TRAF1 (**Figure 3B**), potentially because their effects on other targets counteracted the potential effect on PKN1, although this was not further investigated here.

There was no correlation between TRAF1 levels pre-treatment or of sensitivity to PKN1 inhibition based on Rai stage of the CLL patients. Of 19 CLL patient samples examined for effects of OTSSP167, 16 showed a strong correlation between loss of TRAF1 and gain of caspase 3. Of the 3 outliers, two had very low TRAF1 levels to start with. One CLL patient sample showed an initial small increase in TRAF1 followed by loss of TRAF1 at the concentrations where most cells showed activated caspase 3. Although, IgM mutant status of the donors was not available for these banked CLL samples, the finding that the majority of samples tested responded to a treatment that lowers TRAF1, and that most CLL cells have high TRAF1 levels, suggests the utility of this approach across CLL patient groups. Of note an enrichment for TRAF1^hi^ cells was also observed at suboptimal venetoclax doses for 4/6 donors tested. This might reflect selective loss of TRAF1^lo^ cells, leading to enrichment of the TRAF1^hi^ cells at low inhibitor doses, consistent with an important role for TRAF1 in CLL cell survival and potentially venetoclax resistance. Thus, addition of a TRAF1 inhibitor, via PKN1 inhibition, has the potential to target venetoclax resistance.

While there has been much recent interest and success in targeting antigen receptor signaling and Bcl-2 in B cell malignancies ^5^, to date there has been minimal attention paid to TNFR-mediated survival signaling. TRAF-binding TNFR family members in the tumor microenviroment can contribute key survival signals to CLL cells,^2, 60, 61^ and there is accumulating evidence for the importance of CD40L in promoting drug resistance and contributing to tumor cell survival in the lymphoid microenvironment.^62, 63^ The value of the PKN1-TRAF1 signaling axis as a target for CLL lies in its role downstream of many TNFRSF members and the fact that TNFRSF signaling contributes to NF-κB, MAPK signaling, as well as induction of pS6, an important determinant of cell size and cancer cell fate.^64, 65^ Thus, targeting this pathway could be complementary to drugs that target Bcl-2 family members and/or BCR signaling. In particular, we showed that OTSSP167 synergizes with venetoclax in inducing cell death. Although Mcl-1 inhibitors are being developed to target venetoclax resistance,^66^ these inhibitors do not target all Bcl-2 family members. TNFRSF signaling has also been implicated in inducing other pro-survival Bcl-2 family members, such as Bcl-xL and Bfl-1.^67^ In contrast to Bcl-2 family members, which are important in multiple cell types, TRAF1 expression is limited to activated cells of the immune system. Thus, inhibition of PKN1-TRAF1-dependent TNFRSF signaling to inhibit NF-κB, MAPKs and pS6 might offer improved long-term protection against relapse.

## Data Availability

Drug titration data are included in supplementary tables. Flow cytometry FCS files are available from the authors upon request.

## Author contributions

M.I.E., J.C.L. A.A.A.-S. and T.H.W. designed experiments, analyzed data, and wrote the manuscript. J. C.L., M.I.E. and A.A.A.-S, with help from K.T., S.Z. and A.M., performed experiments. A.A. helped with CLL sample coordination and clinical data. D.U., M.I., MP. and R.A. provided medicinal chemistry expertise and contributed the OICR inhibitor library. M.M. provided samples, clinical data and insight.

## Acknowledgements

We thank Ann McPherson for generating TRAF1-knockdown RAJI cells, Juan Carlos Zúñiga-Pflücker for providing OP9 stromal cells, Dionne White, Joanna Warzyszynska, Nathalie Simard and Janine Charron for assistance with flow cytometry, Birinder Ghumman for technical assistance, and all the patients for participating and donating samples to make this study possible. This research was funded by a Foundation grant from the Canadian Institute of Health Research (FDN-143250) and an Innovation grant from the Canadian Cancer Society Research Institute (2012-701326) to T.H.W.

