## Supplemental Material Table SI and Fig S1-S3 for "The PKN1- TRAF1 signaling axis as a potential new target for chronic lymphocytic leukemia"

**Supplementary Materials contents**

Supplemental Tables

Table S1  page 2

The following supplemental tables are uploaded separately as Excel files.
Table S2. Initial screen of OICR kinase inhibitor library against PKN1.
Table S3. Secondary screen, dose-titrations of inhibitors against PKN1.
Table S4. Extract from Table S3 showing the inhibitors selected for further testing.

Figures S1-S3 with legends page  3-5

### TABLE S1

| | | | | | | FISH | | | | TRAF1 | TRAF1 Reduction | |
|---|---|---|---|---|---|---|---|---|---|---|---|---|
| Patient ID | Age Range | Sex | Vital Status | Rai Stage | CD38+ (%) | del(17p) | del(13q) | del(11q) | Trisomy 12 | dMFI | OTSSP167 | XL-228 |
| 0154 | 60-69 | M | alive | IV | N/A | - | + | + | - | N/D | N/D | N/D |
| 0155 | 70-79 | F | deceased | II | 79 | - | + | - | - | 1214 | equivocal | N/D |
| 0440 | 50-59 | M | alive | IV | 0 | - | - | - | - | 949 | Y | Y |
| 0883 | 40-49 | F | deceased | IV | 32 | - | + | - | + | 937 | Y | N/D |
| 5280 | 40-49 | M | deceased | IV | 1 | N/A | N/A | N/A | N/A | 1261 | Y | N/D |
| 6130 | 60-69 | M | lost follow-up | N/A | 0 | N/A | + | - | - | 2997 | Y | Y |
| 6277 | 50-59 | M | lost follow-up | II | 0 | N/A | N/A | N/A | N/A | 2148 | Y | N/D |
| 8179 | 60-69 | M | alive | I | 0 | N/A | N/A | N/A | N/A | 1163 | Y | N/D |
| 8606 | 60-69 | F | deceased | 0 | 0 | - | + | - | - | 285 | N | N/D |
| 080169 | 50-59 | M | alive | II | 0 | N/A | N/A | N/A | N/A | 1331 | Y | Y |
| 080298 | 30-39 | M | alive | II | 0 | - | - | - | - | 1843 | Y | Y |
| 080299 | 60-69 | M | lost follow-up | IV | 0 | - | + | - | - | 1264 | Y | Y |
| 110250 | 40-49 | M | lost follow-up | N/A | N/A | N/A | N/A | N/A | N/A | 1806 | Y | N/D |
| 130705 | 60-69 | M | deceased | IV | N/A | - | - | - | - | 3018 | Y | N/D |
| 140490 | 70-79 | M | deceased | IV | 43 | - | + | - | - | N/D | N/D | N/D |
| 140515 | 40-49 | M | alive | IV | 0 | - | - | - | - | 1097 | Y | Y |
| 140728 | 60-69 | M | deceased | III | 0 | - | + | - | - | 874 | Y | Y |
| 140729 | 50-59 | F | alive | IV | 0 | - | + | - | - | N/D | N/D | N/D |
| 140790 | 60-69 | F | alive | N/A | 0 | - | + | - | + | 2751 | Y | N/D |
| 140984 | 70-79 | F | deceased | IV | 1 | N/A | N/A | N/A | N/A | N/D | N/D | N/D |
| 150639 | 50-59 | M | alive | I | 60 | - | - | - | - | 1379 | Y | Y |
| 151586 | 80-89 | F | alive | IV | 0 | - | + | - | - | N/D | N/D | N/D |
| 160046 | 70-79 | F | alive | III | 46 | - | - | - | - | 688 | N | N |
| 160421 | 40-49 | M | alive | IV | 0 | N/A | N/A | N/A | N/A | N/A | N/A | N/A |
| 161314 | 60-69 | F | alive | IV | 0 | - | + | - | - | 1517 | Y | Y |

**Supplementary Table 1. Patient characteristics.** F: Female, M: Male, Y: Yes, N: No, N/A: Not available. N/D not determined.

**Supplemental Figures**

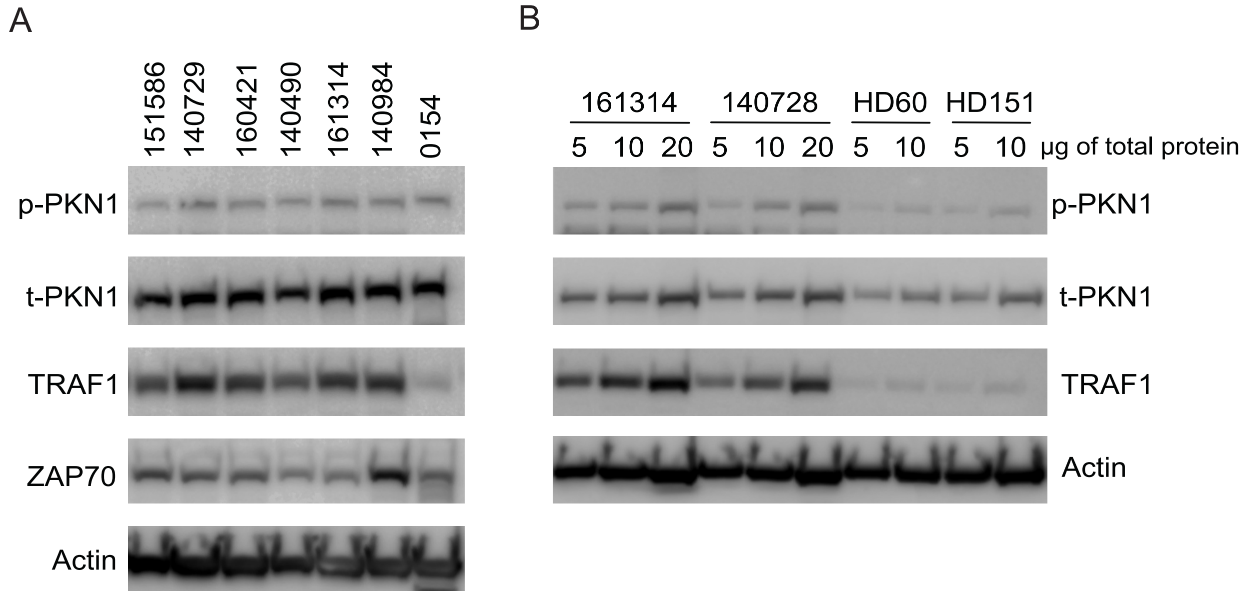


**Figure S1. Phospho-PKN1 and TRAF1 are overexpressed in CLL compared to healthy donors.** (A) 20 µg of whole cell lysates from CLL donor PBMCs were subjected to western blot analysis for Thr774 phospho-PKN1 (p-PKN1), total PKN1 (p-PKN1), TRAF1, ZAP70 or Actin. Each lane represents lysates obtained from individual donors, as labeled. (B) 5, 10 or 20 µg of whole cell lysates from CLL donor PBMCs and 5 or 10 µg of whole cell lysates from isolated B cells from healthy donor PBMC were subjected to western blot analysis for Thr774 phospho-PKN1 (p-PKN1), total PKN1 (p-PKN1), TRAF1 or Actin. Panel A and B show results from a total of 8 CLL patients and 2 healthy donors and are from independent experiments, with donor 161314 repeated in both experiments.

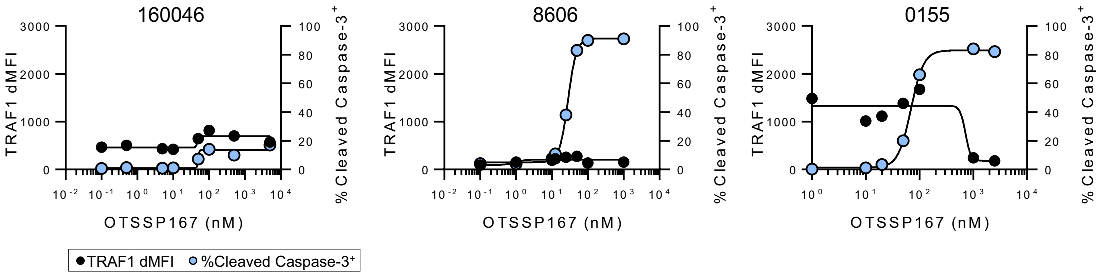


**Figure S2. Response of 3 outliers in TRAF1 response to OTSSP167.** Primary CLL cells were cultured on OP9 stromal cells and treated with OTSSP167 at the indicated concentrations for 24h. Each graph depicts the TRAF1 dMFI and %cleaved caspase-3$^{+}$ CLL cells of one donor. 160046 and 8606 are considered non-responders to OTSSP167 with respect to TRAF1 reduction, 0155 is considered equivocal.

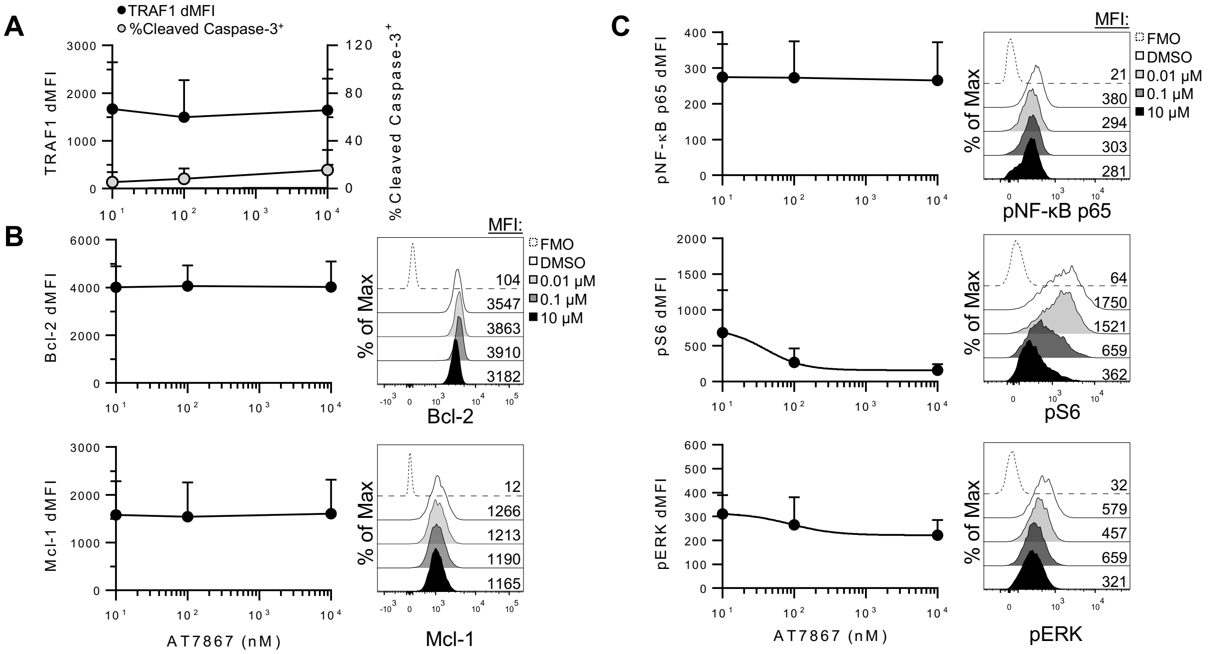


**Figure S3. Effect of AT7867 on TRAF1, cell death, survival and signaling intermediates in primary CLL cells.** Primary CLL cells were cultured on OP9 stromal cells and treated with AT7867 at the indicated concentrations for 24h. Flow cytometry was used to measure: (A) TRAF1 and caspase-3 activation, (B) Bcl-2 and Mcl-1 expression, and (C) pNF-ϰB, pS6 and pERK levels. Line graphs show, for each parameter analyzed, the dMFI or %cleaved caspase-3$^{+}$ cells as mean±SD, n=6 (2 independent experiments pooled). In each panel, representative histograms from one donor show the MFI of AT7867 treated CLL cells and untreated controls.
